# The Effect of Early Hydroxychloroquine-based Therapy in COVID-19 Patients in Ambulatory Care Settings: A Nationwide Prospective Cohort Study

**DOI:** 10.1101/2020.09.09.20184143

**Authors:** Tarek Sulaiman, Abdulrhman Mohana, Laila Alawdah, Nagla Mahmoud, Mustafa Hassanein, Tariq Wani, Amel Alfaifi, Eissa Alenazi, Nashwa Radwan, Nasser AlKhalifah, Ehab Elkady, Manwer Alanazi, Mohammed Alqahtani, Khalid Abdullah, Yousif Yousif, Fouad AboGazalah, Fuad Awwad, Khaled Alabdulkareem, Fahad AlGhofaili, Ahmad AlJedai, Hani Jokhdar, Fahad Alrabiah

## Abstract

**BACKGROUND:** Currently, there is no proven effective therapy nor vaccine for the treatment of SARS-CoV-2. Evidence regarding the potential benefit of early administration of hydroxychloroquine (HCQ) therapy in symptomatic patients with Coronavirus Disease (COVID-19) is not clear.

**METHODS:** This observational prospective cohort study took place in 238 ambulatory fever clinics in Saudi Arabia, which followed the Ministry of Health (MOH) COVID-19 treatment guideline. This guideline included multiple treatment options for COVID-19 based on the best available evidence at the time, among which was Hydroxychloroquine (HCQ). Patients with confirmed COVD-19 (by reverse transcriptase polymerase chain reaction (PCR) test) who presented to these clinics with mild to moderate symptoms during the period from 5-26 June 2020 were included in this study. Our study looked at those who received HCQ-based therapy along with supportive care (SC) and compared them to patients who received SC alone. The primary outcome was hospital admission within 28-days of presentation. The secondary outcome was a composite of intensive care admission (ICU) and/or mortality during the follow-up period. Outcome data were assessed through a follow-up telephonic questionnaire at day 28 and were further verified with national hospitalisation and mortality registries. Multiple logistic regression model was used to control for prespecified confounders.

**RESULTS:** Of the 7,892 symptomatic PCR-confirmed COVID-19 patients who visited the ambulatory fever clinics during the study period, 5,541 had verified clinical outcomes at day 28 (1,817 patients in the HCQ group vs 3,724 in the SC group). At baseline, patients who received HCQ therapy were more likely to be males who did not have hypertension or chronic lung disease compared to the SC group. No major differences were noted regarding other comorbid conditions. All patients were presenting with active complaints; however, the HCQ groups had higher rates of symptoms compared to the SC group (fever: 84% vs 66.3, headache: 49.8 vs 37.4, cough: 44.5 vs 35.6, respectively). Early HCQ-based therapy was associated with a lower hospital admission within 28-days compared to SC alone (9.4% compared to 16.6%, RRR 43%, *p-value* <0.001). The composite outcome of ICU admission and/or mortality at 28-days was also lower in the HCQ group compared to the SC (1.2% compared to 2.6%, RRR 54%, *p-value* 0.001). Adjusting for age, gender, and major comorbid conditions, a multivariate logistic regression model showed a decrease in the odds of hospitalisation in patients who received HCQ compared to SC alone (adjusted OR 0.57 [95% CI 0.47-0.69], *p-value <0.001*). The composite outcome of ICU admission and/or mortality was also lower for the HCQ group compared to the SC group controlling for potential confounders (adjusted OR 0.55 [95% CI 0.34-0.91], *p-value* 0.019).

**CONCLUSION:** Early intervention with HCQ-based therapy in patients with mild to moderate symptoms at presentation is associated with lower adverse clinical outcomes among COVID-19 patients, including hospital admissions, ICU admission, and/or death.

## INTRODUCTION

COVID-19 has rapidly emerged as a pandemic infection that caused significant morbidity and mortality worldwide. Globally, extensive efforts have been made to explore effective and safe therapeutics against the causative virus, SARS-CoV-2 (1). Several medications, including remdesivir, favipiravir, the combination of ribavirin, interferon-beta, and lopinavir-ritonavir, have been suggested based on promising in-vitro results therapeutic experiences from two other coronavirus diseases; severe acute respiratory syndrome and the Middle East respiratory syndrome. However, none of these medications has yet been translated into clinical benefits in treating patients with COVID-19 (2, 3).

Hydroxychloroquine (HCQ), best known as an antimalarial medication, is prominent on the list of potential COVID-19 treatments, owing to its potent antiviral activity against SARS-CoV-2 in in-vitro studies and the results from several trials (4, 5). In-vitro studies show that HCQ blocks COVID-19 infection at a low-micromolar concentration, with a half-maximal effective concentration (EC50) of 1.13 μM and a half-cytotoxic concentration (CC50) greater than 100 μM. The exact mechanism of HCQ’s antiviral activity in HIV is not fully understood, yet several mechanisms have been proposed (7-10). Early theories focused on alterations in post-transcriptional development of the outer HIV surface molecule glycoprotein 120 (gp120), which would render newly formed virions non-infectious (7-12).

To date, studies regarding the efficacy of HCQ, whether alone or in combination with azithromycin, have been contradicting with some pointing towards improved various clinical outcomes (4,5,13-16). In contrast, others failed to demonstrate any benefit (17-21). However, there are major differences amongst these studies in terms of the populations which received HCQ vs a comparator and the timing of initiation of the therapy which may have a significant impact on the variability of these results.

Zinc is a supplement which also has potential antiviral properties that affect the common cold, many of which are due to coronaviruses (22). The combination of HCQ with zinc in the treatment of COVID-19 patients, in an out-or inpatient setting, has been believed to improve the clinical outcome and limit COVID-19 mortality rates, especially if given in early stages of the disease (14). However, evidence regarding the potential therapy of HCQ, whether given alone or in combination with zinc, for COVID-19 patients, is not clear and limited (23) Furthermore, chloroquine and its derivative HCQ may hamper cardiac function at clinically relevant doses, and its safety margin is questionable (20,24). Therefore, further studies are needed to monitor this medication’s safety and benefits.

As part of its response to the COVID-19 pandemic, the Saudi Arabian Ministry of Health (MOH) launched a national fever clinic program to support the acute healthcare system. Healthcare providers at these clinics were managing patients according to a national MOH COVID-19 management guideline which included the option of starting HCQ in addition to the supportive care according to disease severity (25). This study aims to assess the effect of the early use of HCQ in addition to supportive care (SC) compared to supportive care SC alone in patients with confirmed COVID-19 (by Polymerase Chain Reaction (PCR) test) presenting with mild or moderate disease at these ambulatory fever clinics on 28-day adverse clinical outcomes.

## METHODS

### Study setting and design

The national COVID-19 response led by the Ministry of Health (MOH) at Saudi Arabia focused on providing guidance on diagnostic and therapeutic options for COVID-19 as well as improving access to care across the Kingdom. Within that, a comprehensive COVID-19 management guideline was devised by a group of clinical experts according to the best available evidence at the time and was published and periodically reviewed by the MOH (26). This management guideline based the treatment on supportive care therapy in addition to other therapeutics to be considered and included HCQ as a possible option for mild to moderate disease if there was no contraindication.

In line with the national COVID-19 response vision, the MOH also launched a national fever clinic program across all regions of the Kingdom to support the healthcare system. By June 2020, a total of 238 fever clinics were fully operational in assessing patients with symptoms concerning for COVID-19. These fever clinics provided free medical care to all community members regardless of their nationality, insurance status, legal status, and area of residence. The national fever clinic program included screening all patients using an approved national visual triage checklist from the Saudi Center for Disease Control (26), measuring vital signs, detailed assessment by a trained primary care provider, and considering treatment options per the MOH management guideline (25). The fever clinics were designed to care for patients with mild to moderate symptoms, while unstable patients were referred to emergency care services (**appendix.1**). During the selected study period, HCQ was the only available treatment option along with supportive care at these fever clinics. The final decision for starting HCQ therapy in addition to supportive care was based on the individual provider’s discretion after detailed risk assessment (including comorbidity screening, baseline electrocardiogram (ECG), serum electrolytes check) and the shared decision with the patient. Per the ambulatory fever clinic program, patients with baseline abnormal QTc interval or electrolyte imbalances were not prescribed HCQ. Given the overall safety concerns about HCQ therapy in patients above the age of 65 years, the national ambulatory clinic program cautioned providers from prescribing it to this age group. If HCQ was prescribed, patients were required to return for a follow-up visit at day 3 to assess tolerance and to obtain repeat ECG and serum electrolytes to ensure safety. HCQ therapy was discontinued at any time patients reported any medication-related adverse events. All patients who attended these clinics provided consent be enrolled in and allow the use of their clinical data for prospective research purposes at their first visit.

A comprehensive implementation plan was rolled out for this national fever clinic program which included: 1) continuous supply chain of personnel protective equipment, medical devices, and medications; 2) virtual training sessions of 990 primary care providers operating these clinics by an infectious diseases specialist and a senior clinical pharmacist about the clinic program; 3) hotline service to access infectious diseases expertise opinion when needed; 4) standardised ambulatory medication prescription order sets to minimise variability; 5) fever clinics with extended hours of service at 24 hours 7 days a week; 6) extensive media coverage to educate the community about the program; 7) fully equipped call centre to coordinate appointments and answer inquiries around the clock.

This observational prospective cohort study looks at the outcomes of patients presenting to these ambulatory fever clinics during the period between the 5^th^ to 26^th^ of June 2020 who had mild to moderate symptoms and were later confirmed to have COVID-19. All enrolled patients were followed up telephonically at day 28 to record their outcomes (either personally or by a family member).

### Study participants

Symptomatic patients with PCR-confirmed COVID-19 who attended the ambulatory fever clinics during the study period were included in this study. Mild to moderate symptoms included fever (> 38 ⁰C) with or without one or more of the following symptoms: sore throat, cough, diarrhoea, shortness of breath, headache, and myalgia. Patients who were less likely to get HCQ prescriptions were excluded from the study cohort such as paediatrics patients (age < 14 years), pregnant and lactating ladies, patients known to have conductive heart disease, immunocompromising conditions, baseline home oxygen requirement, morbid obesity (BMI ≥ 35), known allergy to HCQ, and glucose-6-phosphate dehydrogenase (G6PD) deficiency.

Study participants were divided into two groups; those who received the SC and those who received HCQ therapy along with the SC. Per the national ambulatory fever clinic program, the SC included symptomatic therapy with zinc sulphate 60 mg once daily for five days, cetirizine 10 mg once daily for 10 days, and paracetamol on an as-needed basis. Those who received HCQ were prescribed a regimen of 400 mg orally twice a day for the first day, followed by 200 mg twice daily for an additional four days according to the MOH management guideline. No dose adjustment was recommended in cases with renal or hepatic impairment.

Patients who had clinical progression or deterioration at day 3 assessment were referred to a hospital setting for management and continued their participation in the study outcome according to their initial assigned group. Study participants who did not show up for their day 3 assessment were excluded.

### Study Outcomes

The primary outcome of interest was hospital admission within 28-days of presentation. The secondary outcome of the study was a composite of ICU admission and/or mortality during the 28-day follow up period.

### Data Collection Tools

A research electronic clinical data collection form (CDF) completed at the national fever clinic program and a follow-up telephone questionnaire done at day 28 were used to collect data about the study participants. Trained primary health care physicians filled out the CDF at day 1 and day 3 assessment visits for each patient per the program requirement. The CDF included patient’s demographics, chronic medical conditions, presenting symptoms, physical exam findings, laboratory results, procedures, and management done at each visit. Data entry officers at the MOH regional Medical affairs entered the data from the CDFs into an advanced national online database. The day 28 telephone questionnaire was conducted by trained personnel who contacted the COVID-19 positive patients or their delegated family members and asked about their clinical outcomes. Outcome data of all the PCR-confirmed COVID-19 patients were also verified with reports from the National disease surveillance database (Health Electronic Surveillance Network, HESN) and the MOH national morbidity & mortality registry. All outcome data were additionally shared with regional Medical Affairs and were further verified with local hospitalisation, ICU, and mortality registries.

### Statistical Analysis

The data were analysed using SPSS^®^ version 25.0. All the data had categorical characteristics, which was described as frequency and percentages. Chi-square test, Fisher exact test, and Crude odds ratio were used to compare symptomatic patients who received HCQ and SC across Socio-demographic background variables and comorbid conditions. Multivariable Logistic regression model was used to assess for primary and secondary outcomes controlling for age, gender, and major comorbidities. The level of significance was considered at P<0.05.

### Ethical consideration

The Saudi Arabian MOH central Institutional Review Board (IRB) approved this observational prospective cohort study, log number: 20-129M. Study enrolment was voluntarily, and all study participants signed an informed consent after receiving a detailed explanation of the research study protocol by their treating physicians. As the study design is purely a prospective observational cohort which followed a predefined population rather than an interventional trial, clinical trial registration was exempted by the MOH Central IRB. The process of prescribing HCQ in COVID-19 followed the national guideline of prescribing recommendation in Saudi Arabia.

## RESULTS

Among 13,592 patients who presented with symptoms to the ambulatory fever clinics during the study period, 7,892 patients had PCR-confirmed COVID-19 of which 5,541 participants responded to the 28-day telephone questionnaire, and their outcome data could be verified with national registries were included in the final analysis. **Figure.1** summarises patient population selection. Among the study participants, almost 33% (n= 1,817) received HCQ in addition to SC while 67.2% (n= 3,724) received the SC only. **Table.1** summarises the socio-demographic and associated comorbidities distribution between the two groups. Significant differences were noted between the groups at baseline, with more males, ages less than 65 years in the HCQ group. There were no significant differences between both groups in terms of overall comorbid conditions except for chronic lung diseases and hypertension with higher percentages among the SC group compared to the HCQ group (1.1% and 9.2% versus 0.4% and 7.2% respectively, *p-value* <0.05). In terms of other administered medications, there was no difference between the two groups in receipt of antibiotics at any point during the study period and follow up; however, the SC group had a higher frequency of receiving steroids after hospitalisation compared to the HCQ group (1.6% vs 0.2%, p-value <0.001).

**Figure. 1:**
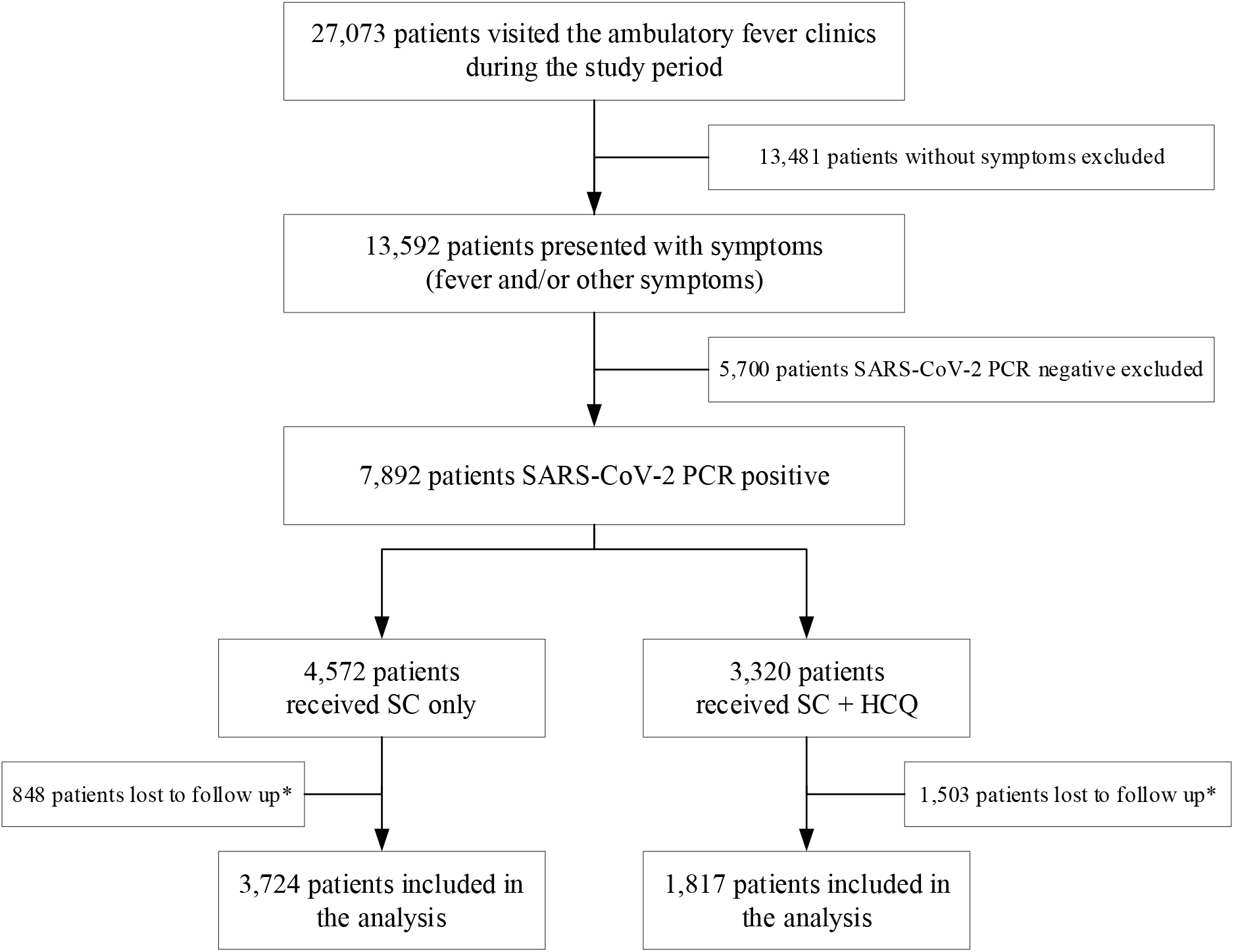
Flow diagram of the cohort selection. **Figure 1 Legend:** Flow diagram of symptomatic COVID-19 patients assessed at the national ambulatory fever clinics in Saudi Arabia during the period from 5-26 June 2020. Outcome recorded at 28-day follow up. *\* The outcome of lost to follow patients were verified with national mortality registry and local hospitalisation and mortality registries and no mortality or hospitalisation were recorded among them*. HCQ = hydroxychloroquine; SC = standard of care; PCR = polymerase chain reaction.

**Table. 1:**
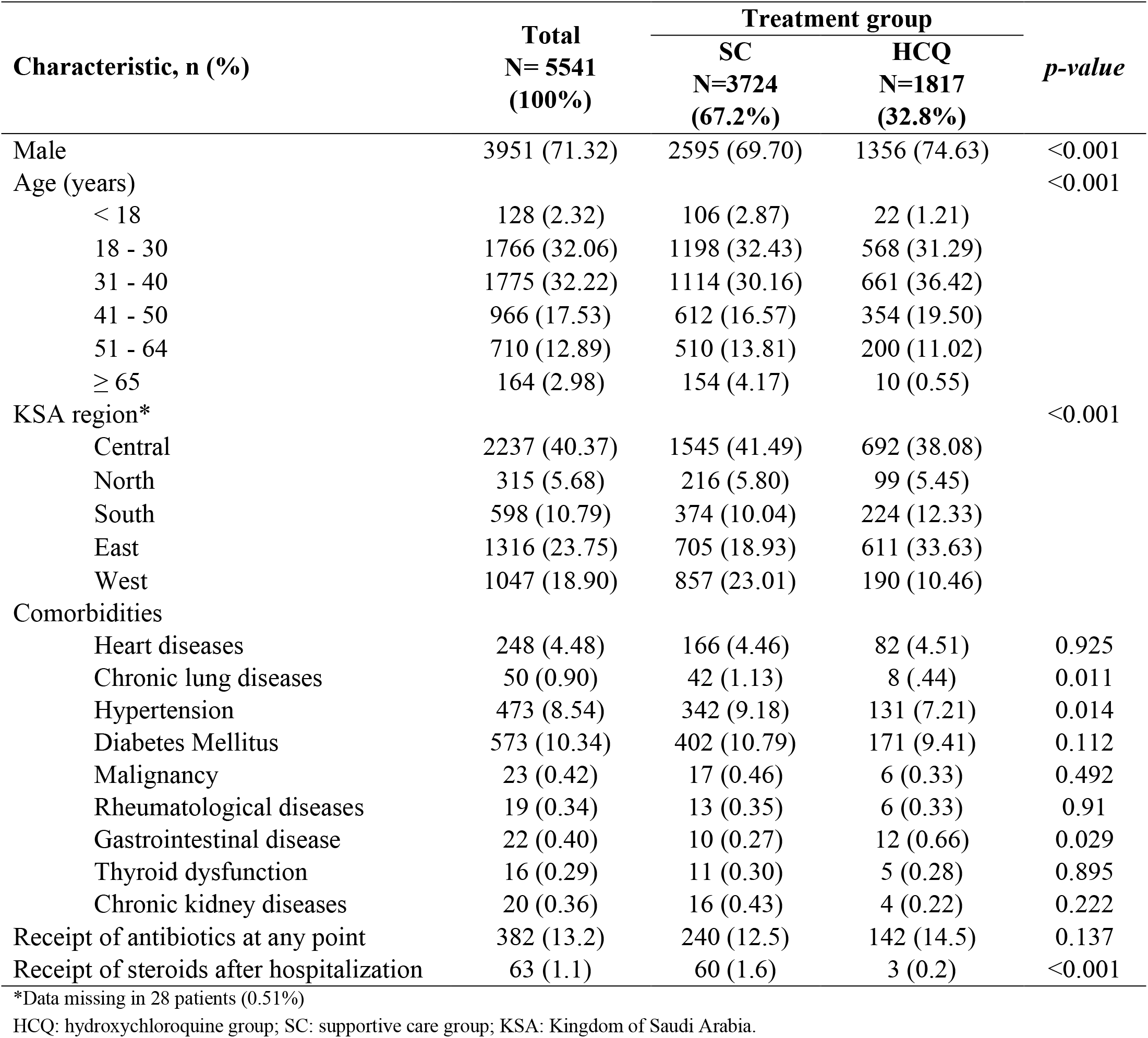
Baseline characteristics of mild-moderately symptomatic COVID-19 Positive patients presenting to the national fever clinic program during the study period.

| Characteristic, n (%) | Total<br>N= 5541<br>(100%) | Treatment group |  | *p-value* |
| --- | --- | --- | --- | --- |
|  |  | SC<br>N=3724<br>(67.2%) | HCQ<br>N=1817<br>(32.8%) |  |
| Male | 3951 (71.32) | 2595 (69.70) | 1356 (74.63) | <0.001 |
| Age (years) |  |  |  | <0.001 |
| < 18 | 128 (2.32) | 106 (2.87) | 22 (1.21) |  |
| 18 - 30 | 1766 (32.06) | 1198 (32.43) | 568 (31.29) |  |
| 31 - 40 | 1775 (32.22) | 1114 (30.16) | 661 (36.42) |  |
| 41 - 50 | 966 (17.53) | 612 (16.57) | 354 (19.50) |  |
| 51 - 64 | 710 (12.89) | 510 (13.81) | 200 (11.02) |  |
| ≥ 65 | 164 (2.98) | 154 (4.17) | 10 (0.55) |  |
| KSA region* |  |  |  | <0.001 |
| Central | 2237 (40.37) | 1545 (41.49) | 692 (38.08) |  |
| North | 315 (5.68) | 216 (5.80) | 99 (5.45) |  |
| South | 598 (10.79) | 374 (10.04) | 224 (12.33) |  |
| East | 1316 (23.75) | 705 (18.93) | 611 (33.63) |  |
| West | 1047 (18.90) | 857 (23.01) | 190 (10.46) |  |
| Comorbidities |  |  |  |  |
| Heart diseases | 248 (4.48) | 166 (4.46) | 82 (4.51) | 0.925 |
| Chronic lung diseases | 50 (0.90) | 42 (1.13) | 8 (.44) | 0.011 |
| Hypertension | 473 (8.54) | 342 (9.18) | 131 (7.21) | 0.014 |
| Diabetes Mellitus | 573 (10.34) | 402 (10.79) | 171 (9.41) | 0.112 |
| Malignancy | 23 (0.42) | 17 (0.46) | 6 (0.33) | 0.492 |
| Rheumatological diseases | 19 (0.34) | 13 (0.35) | 6 (0.33) | 0.91 |
| Gastrointestinal disease | 22 (0.40) | 10 (0.27) | 12 (0.66) | 0.029 |
| Thyroid dysfunction | 16 (0.29) | 11 (0.30) | 5 (0.28) | 0.895 |
| Chronic kidney diseases | 20 (0.36) | 16 (0.43) | 4 (0.22) | 0.222 |
| Receipt of antibiotics at any point | 382 (13.2) | 240 (12.5) | 142 (14.5) | 0.137 |
| Receipt of steroids after hospitalization | 63 (1.1) | 60 (1.6) | 3 (0.2) | <0.001 |

\*Data missing in 28 patients (0.51%)
HCQ: hydroxychloroquine group; SC: supportive care group; KSA: Kingdom of Saudi Arabia.

Per the prespecified inclusion criteria, all patients who were included in the analysis have presented with mild to moderate symptoms concerning for possible COVID-19. Almost all the presenting symptoms were seen in higher percentages among the patients who ended up receiving HCQ therapy compared to the SC alone, most notably: fever (83.91% vs 66.27%), headache (49.78% vs 37.41%), cough (44.54% vs 35.41%), and myalgia (43.65% vs 33.94%) (**Figure.2**).

**Figure 2:**
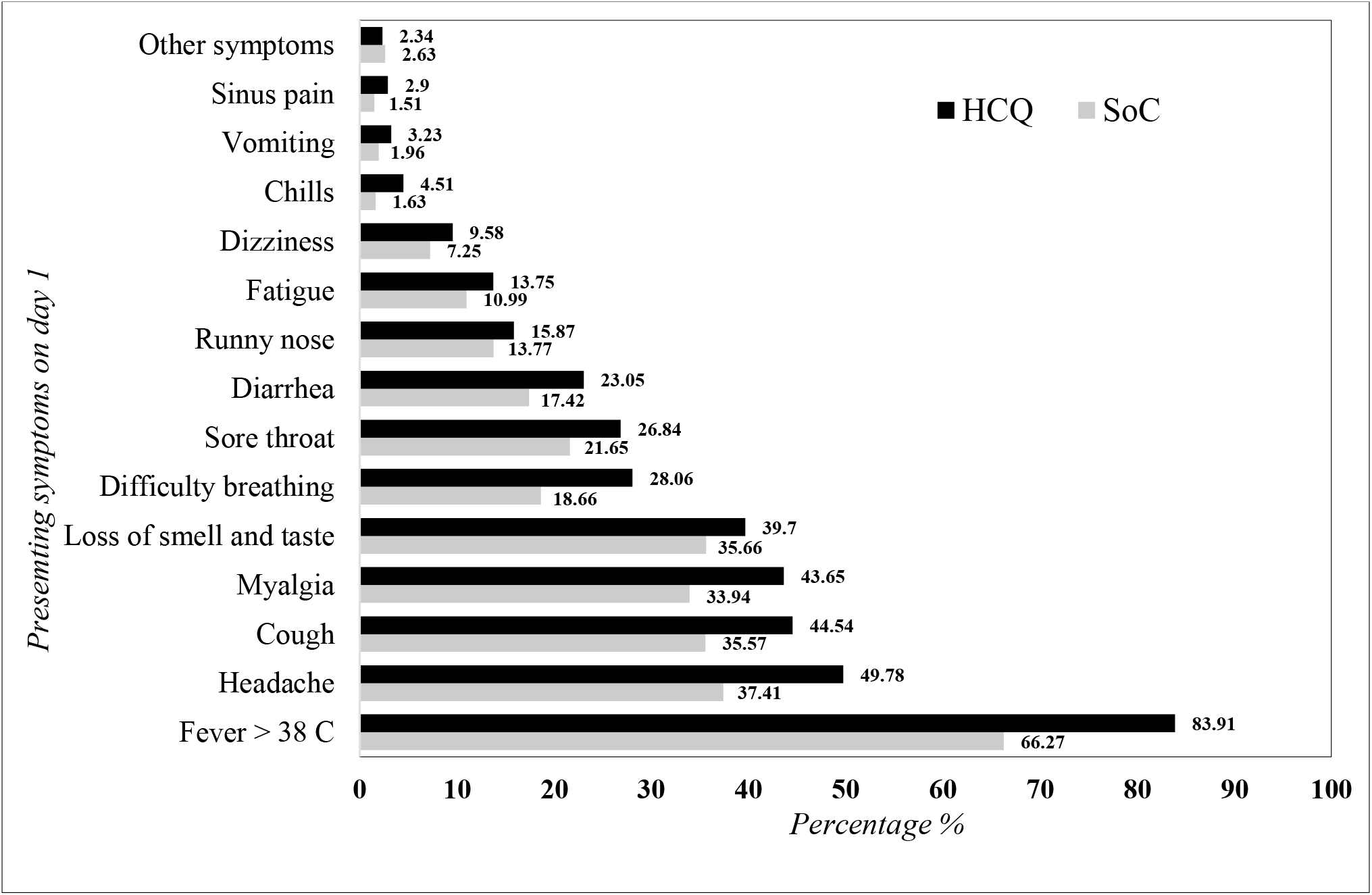
Frequency of COVID-19 symptoms at presentation among patients who received hydroxychloroquine therapy compared to supportive care. **Figure 2 Legend:** Flow diagram of ambulatory symptomatic COVID-19 patients assessed at the national fever clinics in Saudi Arabia during the period from 5-26 June 2020. Outcome recorded at 28-day follow up. HCQ = hydroxychloroquine; SC = supportive care.

The overall hospitalisation rate from disease progression in the study population was 14.2% (N= 788) with significant fewer hospital admissions in the HCQ group compared to the SC (171 (9.36%) vs 617 (16.6%), *p-value* <0.001). This corresponded to a relative risk reduction in hospital admission of 43% among patients who received HCQ compared to the SC (**Table.2**). The rate of ICU admissions and mortality rate were also lower in the HCQ compared to the SC (0.77 vs 1.5 (*p-value* 0.022), and 0.39 vs 1.45 (*p-value* <0.001), respectively). The primary and secondary outcomes of interest were verified with national mortality data and local hospitalisation and mortality registries for all the COVID-19 symptomatic patients at presentation (N= 7,892), and no outcomes were noted in the population which were lost to follow up.

**Table. 2:**
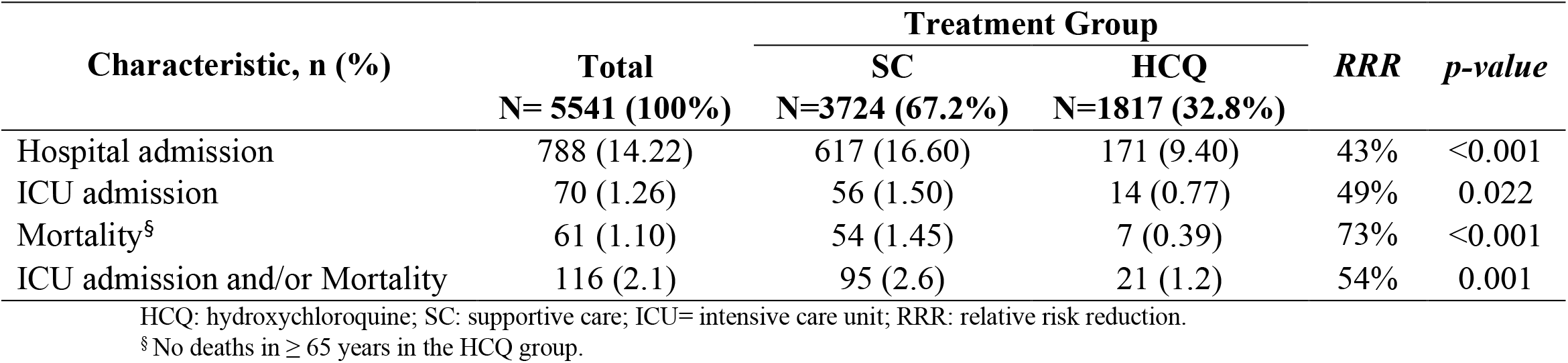
28-days clinical outcomes of COVID-19 positive patients with mild-moderate symptoms who received hydroxychloroquine at presentation to the national fever clinic program compared to those who only received supportive care.

The multivariate logistic regression model shows a significant decrease in the odds of hospitalisation in mild-moderately symptomatic COVID-19 positive patients who received HCQ compared to SC alone, even after adjusting for potential baseline confounders such as age, gender, and major comorbidities (adjusted OR 0.57 [95% CI 0.47-0.69], *p-value <0.001*) (**Table.3**). The composite outcome of ICU admission and/or death was also lower for the HCQ group compared to the SC group controlling for the same prespecified confounders (adjusted OR 0.55 [95% CI 0.34-0.91], *p-value* 0.019). **Table.4** shows the full multivariable logistic regression model.

**Table. 3:** Logistic regression model comparing 28-day clinical outcomes of mild-moderate symptomatic COVID-19 positive patients who received hydroxychloroquine as outpatient compared to supportive care.

| Clinical outcome | Crude OR (95% CI) | Adjusted OR* (95% CI) | *p-value* ** |
| --- | --- | --- | --- |
| Hospital admission | 0.52 (0.44 - 0.63) | 0.57 (0.47 - 0.69) | <0.001 |
| ICU admission | 0.51 (0.28 - 0.92) | 0.63 (0.34 - 1.15) | 0.133 |
| Mortality<sup>§</sup> | 0.26 (0.12 - 0.58) | 0.36 (0.16 - 0.8) | 0.012 |
| ICU admission and/or Mortality | 0.45 (0.28 - 0.72) | 0.55 (0.34 - 0.91) | 0.019 |

\*adjusted for age (reference = age less than 18), male gender, independent comorbidities: (heart disease, chronic lung disease, hypertension, diabetes and other metabolic disorders, chronic kidney disease, malignancy). ICU= intensive care unit.
\*\* for adjusted OR
<sup>§</sup> No deaths in $\geq 65$ years in the HCQ group.

**Table. 4:** Detailed logistic regression model of clinical outcomes of mild-moderate symptomatic COVID-19 positive patients at 28-days who received hydroxychloroquine as outpatient compared to supportive care.

| Covariate | Adjusted OR (95% CI) | *p-value* |
| --- | --- | --- |
| **Hospital admission** | 0.57 (0.47 - 0.69)* | <0.001 |
| **ICU admission and/or mortality** | 0.55 (0.34 - 0.91)* | 0.019 |
| **Age (years)** |  |  |
| < 18 | Ref |  |
| 18 - 30 | 2.22 (1.38 - 3.55) | <0.001 |
| 31 - 40 | 2.77 (1.73 - 4.43) | <0.001 |
| 41 - 50 | 2.74 (1.7 - 4.43) | <0.001 |
| 51 - 64 | 1.91 (1.17 - 3.14) | 0.007 |
| $\geq 65$ | 0.33 (0.15 - 0.73) | 0.011 |
| **Gender (male)** | 1.23 (1.08 - 1.4) | 0.002 |
| **Comorbidities** |  |  |
| Heart disease | 1.12 (0.85 - 1.48) | 0.429 |
| Hypertension | 1 (0.79 - 1.27) | 0.973 |
| Chronic lung disease | 0.56 (0.26 - 1.21) | 0.141 |
| Diabetes mellitus | 1.14 (0.92 - 1.41) | 0.244 |
| Chronic kidney disease | 0.81 (0.26 - 2.53) | 0.715 |
| Malignancy | 0.77 (0.3 - 2) | 0.594 |

ICU= intensive care unit.
\*The values presented represent the result of independent models which were performed on each outcome separately including the same listed covariates. The adjusted ORs and 95% CI of the age, gender, and comorbidities were the same in both models thus presented once.

## DISCUSSION

Our study is a large observational nationwide cohort of PCR-confirmed COVID-19 patients who presented with mild and moderate symptoms to ambulatory fever clinics and were managed according to a national management guideline which included the prescription of HCQ at an early stage of the disease (25). We describe what happened in real-world clinical practice where the decision to start HCQ therapy was based on the physician risk assessment and the shared decision with the patient which allows assessing the benefit of such intervention if it to be deployed on a population level. Despite the seen differences in the baseline characteristics between the patients who received HCQ and those who received the SC alone, the multivariate logistic regression model that controls for patient-specific prespecified potential confounders shows a lower odds of adverse clinical outcomes, namely, hospitalisation and ICU admission and/or mortality within 28-days of the presentation by 43% and 45% respectively. The decision to start treatment did not differentiate between a specific symptom or combination of symptoms and many patients presented with a group of symptoms thus given the dependent nature of this variable; it was not included in the final multivariable model. As the study protocol did not interfere with the acute care management of the study participants who were hospitalised, it is reasonable to believe that ICU admission criteria would vary between different hospital settings. Nonetheless, there was a trend towards lower ICU admissions in the HCQ group. As the mortality rate in Saudi Arabia is considered low compared to other nations (26, 27), to ensure the stability of the multivariate logistic model, the mortality outcome was looked at as a composite of ICU admissions and/or mortality which reached clinical significance while controlling for the prespecified confounders favouring the effect of early intervention with HCQ.

Per the national ambulatory fever clinic program, at the specified study period, steroid therapy was not advised for the sake of COVID-19 infection per se and was mainly prescribed as indicated, if any. The fact that the receipt of steroid after hospitalisation was significantly higher at the SC group is reassuring that the observed result represents the effect of the early intervention with HCQ rather than the possible confounding effect of early steroid therapy, however, since complete data about steroid prescription at presentation is lacking, this cannot be firmly concluded. Finally, the safety of HCQ therapy in our cohort is described in detail elsewhere, and it was shown to be a tolerable medication with minimum side effects (data submitted for publication by Mohana et al.).

The previously published observational studies which failed to translate the in-vitro mechanistic benefit of HCQ on clinical outcomes mainly introduced the therapy on hospitalised patients (17-21). However, recent large cohort studies showed significantly improved outcomes in patients who received HCQ early during hospitalisation (4,15). This spiked the interest in testing the effect of early administration of HCQ therapy during the initial viral replication phase prior to the progression to the hyperimmune response phase owing to its variable antiviral properties (28). While an Italian multicentre, open-label, randomised controlled trial did not show benefit of early administration of HCQ therapy to mildly symptomatic young adults (29), other retrospective studies showed a promising benefit of early HCQ treatment in modifying the overall outcome of COVID-19 whether or not it was associated with azithromycin (30,31). Our study further supports these later findings and suggests a possible benefit of this early intervention in preventing adverse clinical outcomes on a population level.

Although our study included a large cohort of symptomatic COVID-19 participants, we acknowledge that it has several limitations. The population represented in the dataset analysed is relatively young with a limited number of patients who were above the age of 65 years based on the cautionary measure taken by the national ambulatory fever clinic program. Although the multivariable model adjusts for this age group, given the small numbers of patients in this stratum, we caution from generalising the results to this age-group. Furthermore, the study took place in all regions of the Kingdom during the pandemic, which imposed some logistic challenges leading to losing the follow up of many patients in both treatment groups. To overcome this anticipated challenge, the study protocol was designed with an additional verification process to ensure capturing all hard outcome data from reliable national registries. As this verification process was non-differential to the initial treatment group allocation, and the fact that the sample size of the cohort is considered large, we believe that the overall results are valid.

## CONCLUSION

Although our study population were young and with a relatively low incidence of comorbidities in both treatment groups, early intervention HCQ-based therapy in an ambulatory setting in mild to moderate COVID-19 patients was associated with lower odds of hospitalisation and ICU admission and/or death. Additional large randomised controlled trials are recommended to further support this conclusion, particularly in older populations.

## Data Availability

All data available upon request

---

**Appendix.1:**
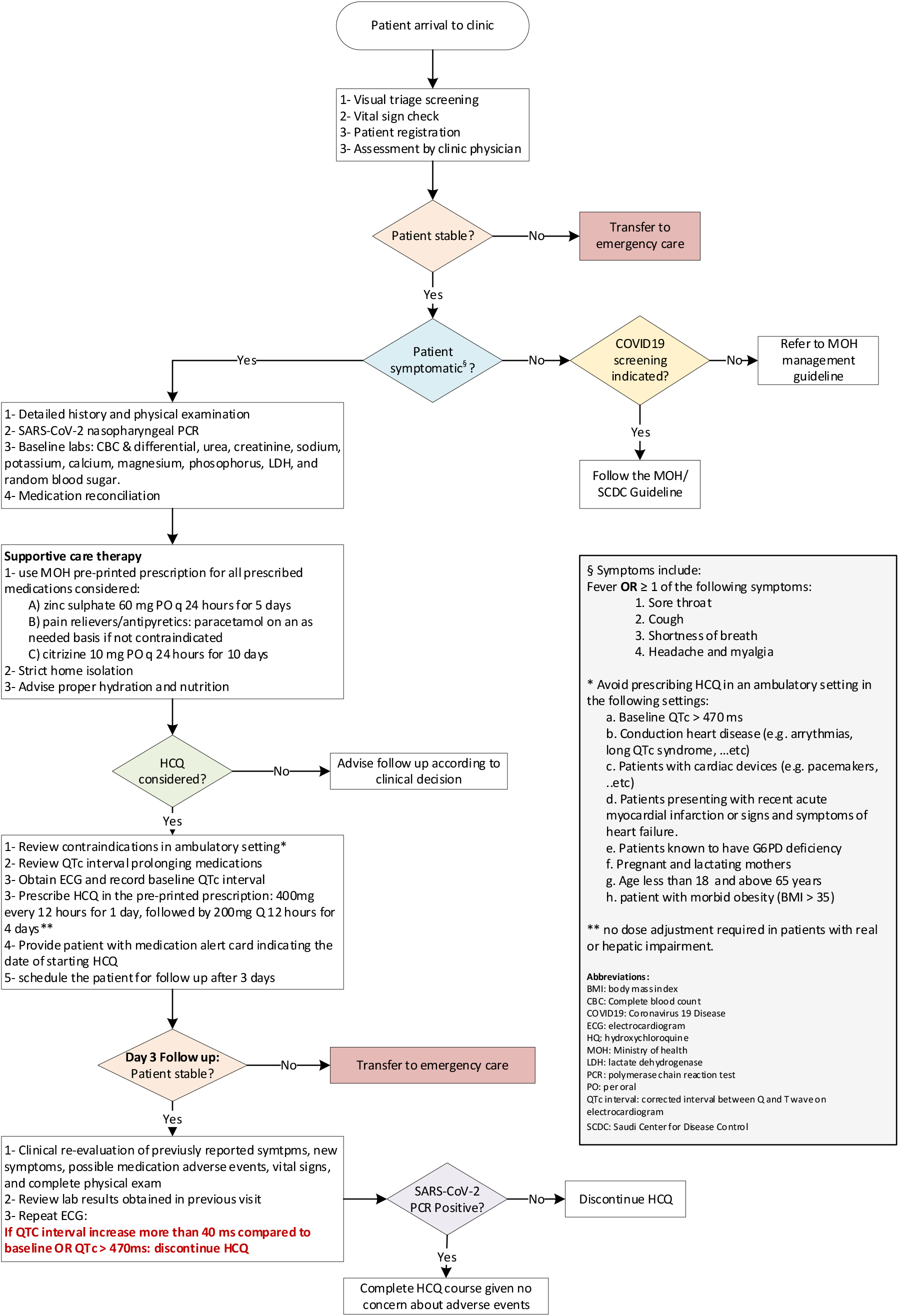
The Saudi Arabian Ministry of Health ambulatory fever clinic program recommendation for patients presenting with mild to moderate symptoms during the COVID-19 pandemic.

